# Pay-it-forward strategy reduced HPV vaccine delay and increased uptake among catch-up age girls: A randomized clinical trial

**DOI:** 10.1101/2025.01.16.25320655

**Authors:** Jing Li, Yifan Li, Chuanyu Qi, Yu He, Haidong Lu, Yewei Xie, Jason J. Ong, Yajiao Lu, Ying Yang, Fan Yang, Heng Du, Wenfeng Gong, Fei Zou, Heidi J. Larson, Mark Jit, Leesa Lin, Jennifer S. Smith, Elvin H. Geng, Dong Xu, Weiming Tang, Shenglan Tang, Joseph D. Tucker, Dan Wu

## Abstract

**Background:** Catch-up HPV vaccination is challenging in many low and middle-income countries (LMICs). Pay-it-forward offers an individual a subsidized vaccine, then an opportunity to donate to help others access vaccinations. Our randomized control trial assessed the effectiveness of pay-it-forward in improving HPV vaccination among girls aged 15-18 years in China.

**Methods and findings:** Eligible participants were randomly assigned to either the pay-it-forward arm or standard-of-care arm (self-paid vaccination). The primary outcome was the first-dose HPV vaccination rate, verified against clinical records. Among 321 participants enrolled, most caregivers were female (80.1%). In the pay-it-forward arm, 55 of 161 (34.2%) girls received the HPV vaccine, compared with 28 of 160 (17.5%) girls in the standard-of-care arm (adjusted proportion difference = 17.9%, 95% CI: 8.7, 27.0, P<0.001). Among 55 girls in the pay-it-forward arm who received the vaccination, 37 (67.3%) wrote a postcard message, and 39 (70.9%) of their caregivers donated to support future girls. The financial cost per person vaccinated was $294 in the standard-of-care arm and $230 in the pay-it-forward arm.

**Conclusions:** The pro-social pay-it-forward strategy was effective to increase catch-up HPV vaccination among teenage girls with comparable costs.

**Trial Registration:** ChiCTR2200055542

## Introduction

Catch-up HPV vaccination is challenging, and barriers exist in many low and middle-income countries (LMICs). Despite the clear benefits of free HPV vaccines, many health systems still impose fees. The cost of a single dose of HPV vaccine is comparable to a young woman’s earnings over several months in LMICs. Such costs hinder vaccine uptake, especially among the most vulnerable,[1] erode trust in healthcare[2], and impede universal health coverage goals. Providing free HPV vaccines could accelerate the achievement of HPV vaccine uptake goals. However, countries where free HPV vaccination is available have suffered from low uptake,[3] indicating other barriers in addition to fees. Lack of awareness and vaccine hesitancy may be important drivers in these settings.[4] Misinformation can also undermine confidence in vaccines. Public engagement is essential for tackling such barriers and gaining community acceptance of HPV vaccination,[5, 6] but effective community-engaged programs are limited.[7]

The Chinese government has started local programs to expand subsidized HPV vaccines among 13-14 years old girls in China, but left out older girls, and catch-up vaccination rates among adolescent girls were low (<15%).[8, 9] To achieve the cervical cancer elimination goal, catch-up age groups should also be included.[10] Budget constraints prohibit government’s extension to older age groups, and women are unable or reluctant to self-pay for the widely available vaccine.

Earlier HPV vaccination is recommended for several benefits, i.e., better protection effects, fewer doses and lower costs.[11] However, vaccine delay, defined as receiving the vaccination after the recommended age,[12] remains a significant barrier among many financially capable individuals.[13] Evidence shows that over 61% of Chinese caregivers had a high level of willingness to vaccinate their girls,[14] but 80%[15] opt to postpone vaccination at a later age due to poor awareness, the belief that their girls are sexually inactive with a low risk of exposure.[13] A novel community-engaged solution is urgently needed to address the above financial and perception-related challenges.

Pay-it-forward, an approach that combines incentive and priming nudges, may provide such an opportunity. It gives a person an incentive nudge (e.g., subsidized vaccine) and a kindness-based priming nudge (e.g., community handwritten postcard), and then offers the person an opportunity to give back to the community by donating money and/or creating HPV vaccine promotion messages (interpersonal affecting nudging) (appendix figure 1, p 2).[16] Pay-it-forward builds on the theory of upstream reciprocity (i.e., individuals helped by someone are more likely to help others).[17] Our earlier research demonstrated that pay-it-forward increased health services uptake among priority populations by generating community engagement and enhancing participant confidence in vaccine importance, safety, and effectiveness.[18–20] In this two-arm randomized controlled trial, we aimed to evaluate whether the community-engaged pay-it-forward strategy would reduce vaccine delay behavior and increase earlier HPV vaccine uptake against the standard self-paid vaccination among girls aged 15-18 years in Chengdu, western China where a lower HPV vaccine uptake was observed (8.6%) as compared to other regions. [8, 9]

**Fig 1.**
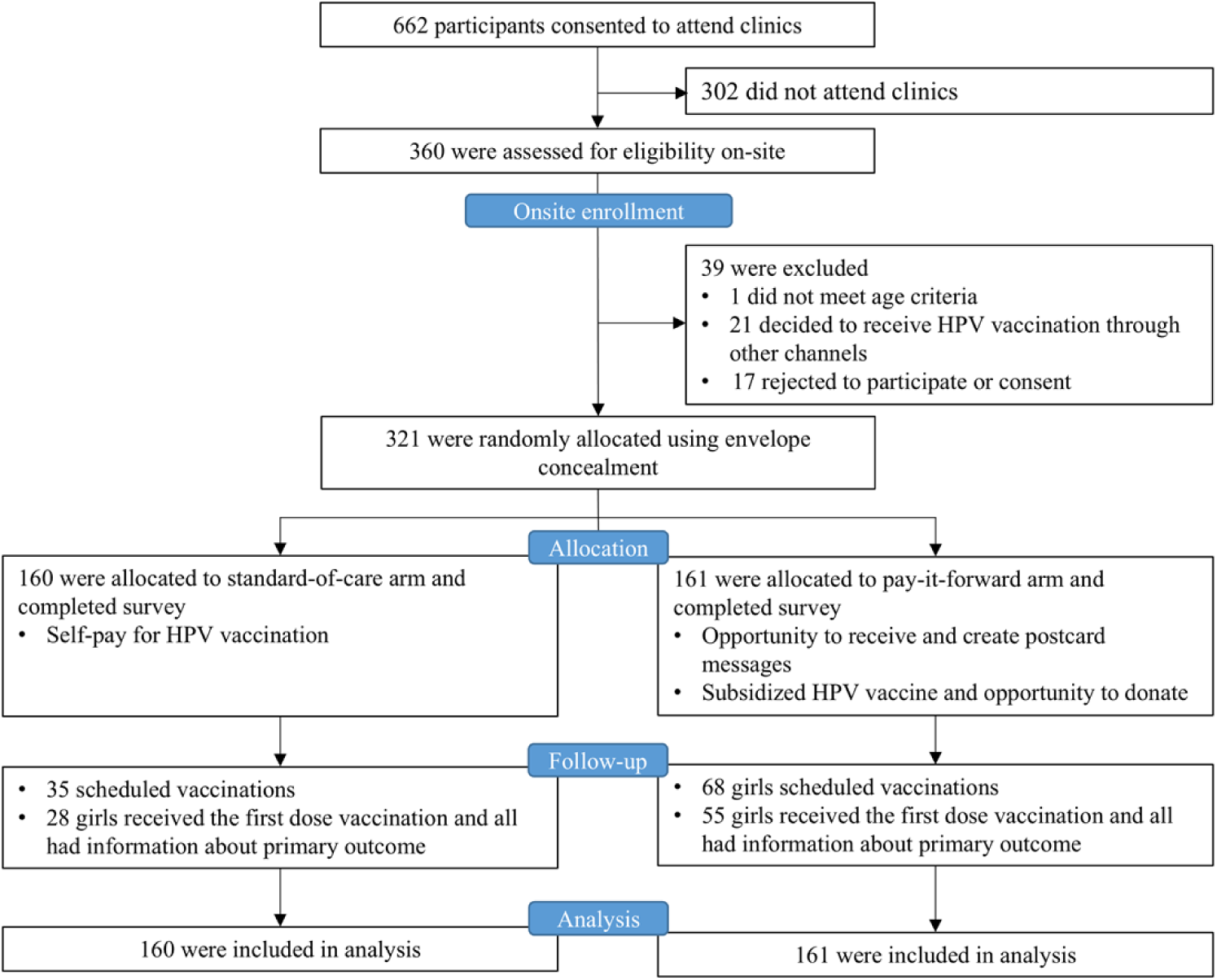
Trial profile and CONSORT diagram.

## Methods

### Study design

This was a two-arm randomized controlled trial (RCT) conducted in Chengdu, Sichuan Province, China. Chengdu had a population of 21 million people in 2022. Chengdu is located in the less developed western part of China and has substantial economic inequality, with both more and less developed areas. The trial was organized at four community health centers that provide routine vaccination services. The protocol has been published (appendix protocol, pp 12-26).[21] Four community health centers were selected because they had the following: essential infrastructure for providing HPV vaccination services, including personnel; and stable HPV vaccine supply. The four sites by relative level of individual disposable income in 2021 were Site A (most developed urban area), Site B (higher middle-income suburban area), Site C (lower middle-income suburban area), and Site D (least developed area).[22] The Ethics Committee of West China Fourth Hospital and West China School of Public Health approved the study.(Gwll2021057/ Gwll2023125). This trial had been registered on Chinese Clinical Trial Registry and the WHO International Clinical Trials Registry Platform (ChiCTR2200055542). The trial URL is https://trialsearch.who.int/Trial2.aspx?TrialID=ChiCTR2200055542. We used CONSORT guidelines[23] for reporting this RCT trial (appendix pp 39-41).

### Participant recruitment and screening

We used the simple random sampling method to select girls. The complete name list of adolescent girls aged 15–18 years in the study area was obtained from the neighborhood committee via community health center staff. This list included all girls of this age range living in the neighborhood who registered their names and contacts of caregivers at the community health center to receive relevant health services, including vaccines. We assigned a random number to each individual using Microsoft Excel. Individuals were sorted by random numbers from the smallest to the largest, and we selected potential participants from the top of the list for telephone recruitment until we reached the sample size of 80 eligible girls at each study site.

Telephone recruitment included a brief introduction to the overall study purpose, procedures, and associated benefits and risks (appendix script, p 11). Inclusion criteria for participants were 1) adolescent girls aged between 15 and 18 years with a caregiver, 2) living in the areas that the chosen community health centers serve, 3) no history of HPV vaccination, 4) no known vaccine allergy. Caregivers of girls confirmed eligibility and were then invited with their girls to the community health center. All caregivers signed a written informed consent form and girls provided assent. Recruitment occurred over a prolonged eight-month period because of COVID-19 lockdowns and community re-direction of vaccine services to prioritize COVID-19 vaccination. Site D recruitment coincided with the relaxation of COVID-19 policies when many clinics were overwhelmed with COVID-19 and vaccine sites were short staffed (appendix figure 2, p 2).

**Fig 2.**
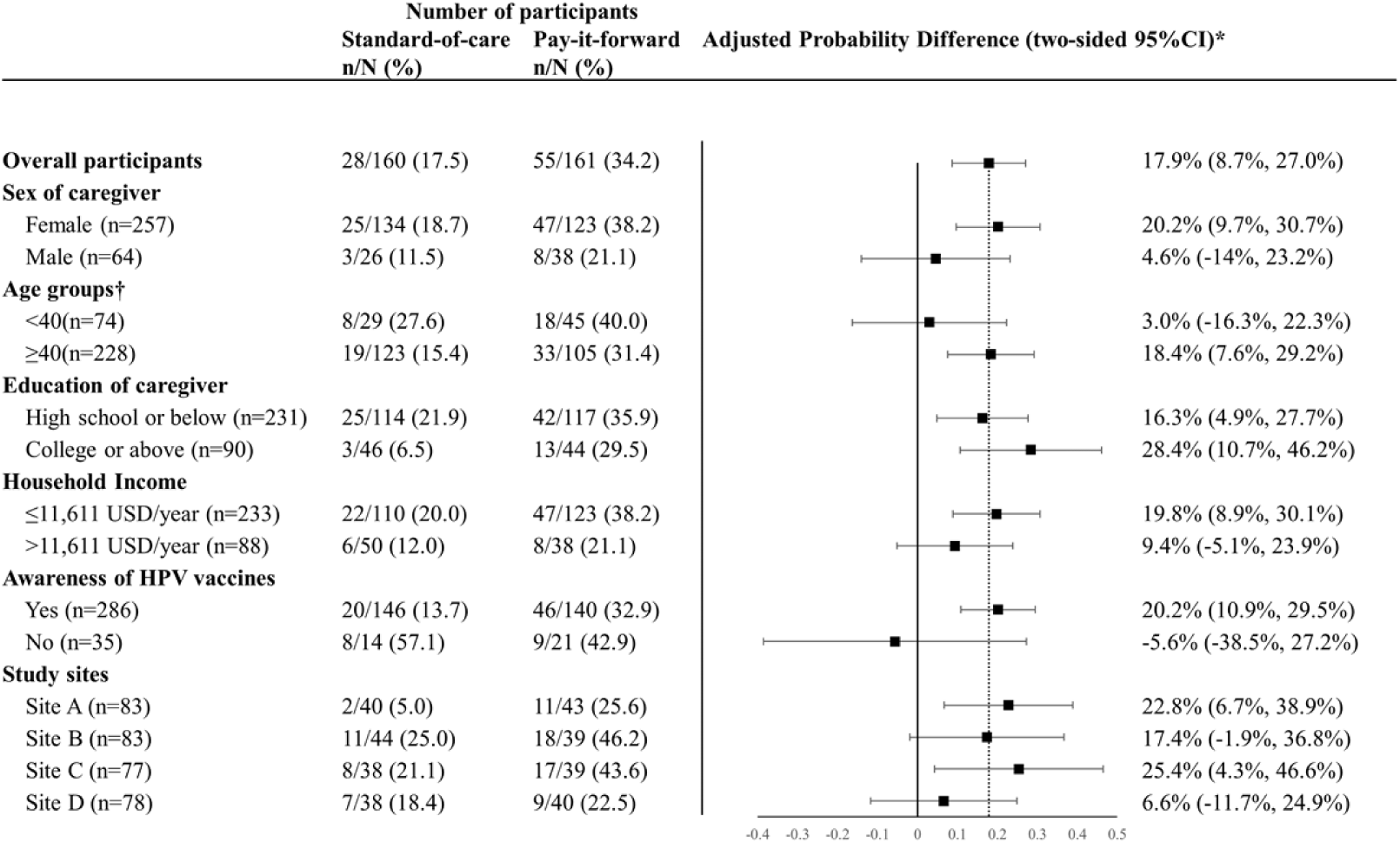
Generalized linear models to compare HPV vaccine uptake rates of two arms. 1USD=6.89RMB. HPV = human papilloma virus. *The proportional differences were adjusted for study sites, household income level, education level and marital status and sex of the caregiver. †There are missing data of the age of caregiver.

### Randomization and masking

Simple randomization of participants was used. An independent statistician prepared the computer-generated randomization list, and we used the sealed envelope method to randomize eligible participants (caregiver-girl pairs) into standard-of-care and pay-it-forward arms at a 1:1 ratio within each center, based on the order in which they visited the sites. An individual-based randomization approach for efficacy evaluation of a newly developed strategy was appropriate.[24] The community health staff coordinator, physicians who prescribed HPV vaccines, the outcome assessors, and data analysts were blinded to intervention allocation. Additionally, the following techniques were adopted to ensure the concealment of allocation in this trial: the process of generating the sequence of random digits and assembling envelopes was confidential to all on-site research staff who recruited participants; the allocated intervention was concealed from both participants and research staff onsite prior to assignment until they opened the envelope.

### Procedures

Caregiver-girl pairs in the standard-of-care arm received a routine educational pamphlet used in clinics and were informed of the prices of available HPV vaccines. They had to pay out of pocket at the standard price if they wanted to receive HPV vaccination.

In addition to the routine educational pamphlet, caregiver-girl pairs in the pay-it-forward arm received community co-created postcard messages (appendix figure 3, p 2-3), subsidized HPV vaccination, and the opportunity to voluntarily donate towards someone else’s vaccine dose and/or write postcard messages. Participants in the pay-it-forward arm were told the market prices to receive available HPV vaccines, and that previous participants had donated RMB 329 (47.7 USD, equal the price of the first dose of domestic HPV vaccination) towards the price, along with postcard messages for them. They were also told that other adolescent girls from their neighborhood cared about them. We used a series of community engagement methods, including stakeholder meetings (n=18), participant interviews and focus group discussions (n=21), postcard messages co-designed with college students (n=4), to adapt and refine the intervention components taking population characteristics and contextual factors into consideration (appendix p 10: community engagement section). Participants in both arms were given a brief introduction to the project, pamphlet information about cervical cancer, HPV vaccination details, and the price of HPV vaccine (appendix figure 4, pp 4-8).

**Fig 3.**
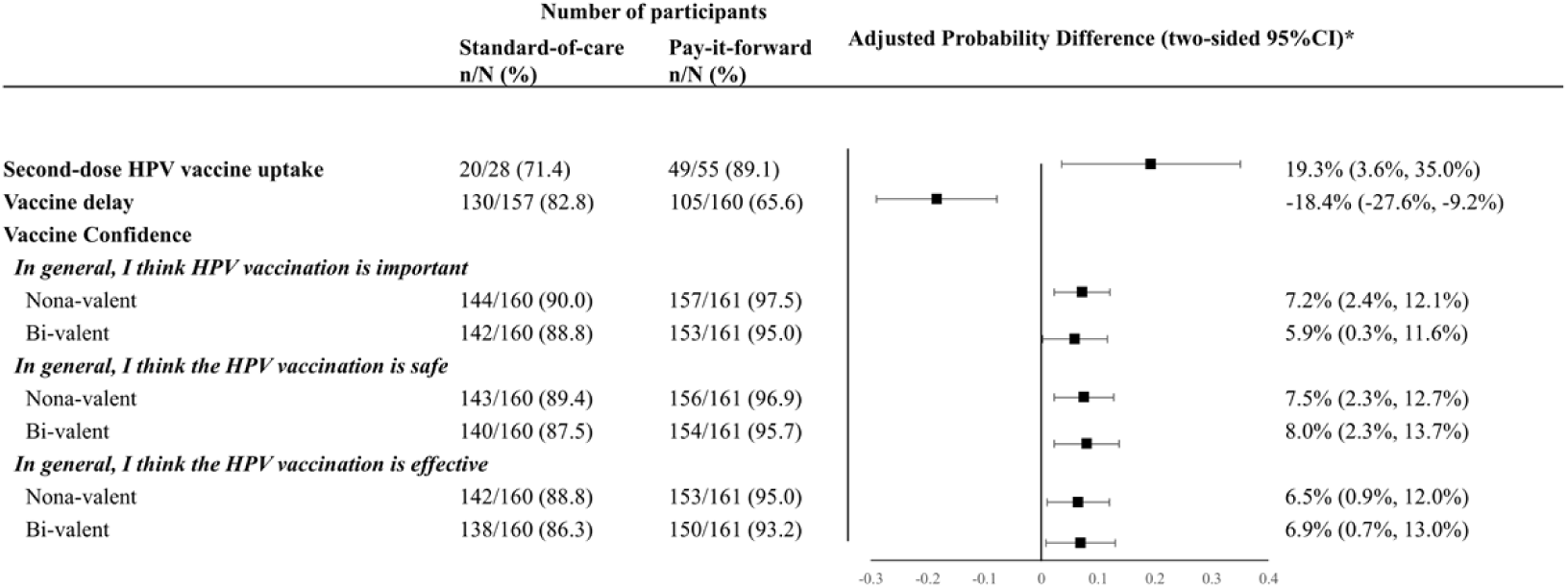
Generalized linear models to compare second-dose HPV vaccine uptake, vaccine delay and vaccine confidence rates of two arms. *The proportional differences were adjusted for study sites, household income level, education level and marital status and sex of the caregiver.

Caregivers who agreed to have their daughter receive a vaccine made an appointment for their daughter to receive vaccination within three months. In the pay-it-forward arm, girls received subsidized vaccination and handwritten postcards and they had an opportunity to create their own postcard messages for others, while caregivers were asked about donation during the half-hour post-vaccine monitoring period. A donation/postcard collection box was provided on-site, and QR code was provided to those who preferred online donation. The donation was voluntary and anonymous. Project staff left participants after bringing them the postcards, and they were unaware of participants’ donation behavior and donation amount.

We used information tracking sheet to collect data about vaccine uptake and adverse events within 24 hours, which were verified against clinical records. All participating caregivers completed an online questionnaire (appendix 35-38) on demographic information and vaccine attitudes. The number of participants who donated, and the donation amount in the pay-it-forward arm were collected. The cost data were obtained from expense records and by asking local health staff.

### Outcomes

The primary outcome of the study was first-dose HPV vaccine uptake within three months after receiving the intervention and ascertained by clinical records. The four community health centers had a stable supply of bivalent HPV vaccine (Cecloin) only during the study period.

Secondary outcomes included second-dose HPV vaccine uptake, number of pay-it-forward participants who donated, donation amounts received, the costs for implementing the intervention and cost per person vaccinated from the healthcare provider perspective, vaccine confidence, and vaccine delay.[12] Vaccine confidence was measured using standard survey items adapted to assess HPV vaccine confidence in China.[25] Vaccine delay refers to individuals have positive intentions to vaccinate but fail to act.[26] The measurement of vaccine delay was adapted from a study evaluating COVID-19 vaccine delay.[27] We asked participants’ willingness to receive the vaccine, and the actual vaccination was verified using clinical records after the 3-month follow-up period.

### Statistical analysis

Details about the sample size calculation are available elsewhere.[21] In short, based on current vaccination rates of <15%[8, 9] and the proportion difference of 16% between the two arms in our pilot study,[21] we estimated that the outcome vaccination rate of the standard-of-care arm was 20% due to the provision of routine educational pamphlet, and 36% in the pay-it-forward arm. Assuming a two-sided Z-Test with unpooled variance, a sample size of 240 (or 120 in each arm) can achieve 80% power in detecting differences between the two arms with a significance level of 0.05. Given a non-response and/or dropout rate of 10%, 133 participants for each arm were needed. To allow sub-analyses on secondary outcomes and sub-group analyses, we increased the sample size by 20% to 160 in each arm.

We followed a preregistered analysis plan and appendix page 255-26 clarified amendments. Descriptive analyses of socio-demographic characteristics and vaccination rates by study arms were conducted. The pay-it-forward and standard-of-care arms’ uptake rates were compared using chi-square tests or Fisher’s exact test. Crude and adjusted proportional differences in HPV vaccine uptake comparing the pay-it-forward arm with standard-of-care arm were estimated using generalized linear models with Gaussian distribution and identity link function, and the corresponding 95% confidence intervals were calculated using sandwich variance estimator.[28] The adjusted proportional differences accounted for study sites, household income level, education level, marital status and sex of the caregiver. The secondary outcomes - vaccine confidence and vaccine delay - were summarized using descriptive statistics. Similarly, comparisons of these secondary outcomes between the two arms were also assessed using generalized linear models with Gaussian distribution and identity link function. Difference in donation amount between different sites in the pay-it-forward arm was assessed using independent-Samples Kruskal-Wallis Test.

We employed a micro-costing method to evaluate the financial cost of implementing HPV vaccination under the two intervention strategies from the health provider’s perspective (Sichuan Department of Health). We tracked all resources used during the experiment and classified cost items. The collected cost data and costs-related results are in the appendix pp 11-12. We presented the total financial cost for each group, the cost per person vaccinated, and the donation in the pay-it-forward group. All expenses are expressed in 2022 USD using OANDA currency conversions (1 USD = 6.89 RMB, year of 2022).

Subgroup analyses were pre-specified to determine whether the effects on vaccine uptake varied by (1) household income levels (≤11,611 USD/year or > 11,611 USD/year). This threshold was determined based on the mean household income in rural Sichuan Province in 2021;[29] (2) study sites A, B, C, D; (3) sex of the participating caregiver (female or male); (4) highest educational attainment of the participating caregiver (college and above, or high school and below); (5) age of caregivers (<40 or ≥40 years old); (6) awareness of HPV vaccine (yes or no). Both crude and adjusted proportional differences in HPV vaccine uptake comparing the pay-it-forward arm with standard-of-care arm for each subgroup were reported. Data were exported to R Statistical Software (version 4.2.1; R Foundation for Statistical Computing, Vienna, Austria; appendix data analysis code, pp 27-34) for analysis. We conducted the cost analysis in Excel 2019 (Microsoft, USA).

The leading academic research organization (Nanjing Medical University, NMU) monitored the study. No data safety monitoring committee was appointed. This study was registered with ChiCTR (ChiCTR2200055542), and is now closed for recruitment.

## Results

From July 6, 2022, through June 9, 2023, we successfully invited a total of 662 eligible caregiver/daughter pairs to attend clinics, and 360 were present for eligibility screening at clinics (clinic response rate = 54%) (Fig 1). Overall, 39 were further excluded due to age ineligibility (n=1), decision to receive HPV vaccination via other channels (n=21) or refuse to consent to participate (n=17), and 321 were included in the final data analysis: 160 were assigned to the standard-of-care arm, and 161 were assigned to the pay-it-forward arm.

The baseline characteristics were similar with no statistical difference between the two arms (table 1). Most caregivers were female (80.1%), married (80.4%), reported a household income level of ≤11,611 USD/year (72.6%), and had an educational background of high school or less (72.0%). Most participants (90.7%) reported no family history of HPV or cervical cancer diagnosis.

**Table 1.** Characteristics of the girls 15-18 years old and their caregivers in Chengdu, China, 2022-2023, n=321*.

|  | Standard-of-care<br>(n=160) | Pay-it-forward<br>(n=161) | Total<br>(n=321) |
| --- | --- | --- | --- |
| Age (girl) — years | 16.6±0.9 | 16.6±1.0 | 16.6±1.0 |
| Age (caregiver) — years | 44.4±8.6 | 43.0±8.5 | 43.7±8.6 |
| Sex of caregiver |  |  |  |
| Male | 26(16.3) | 38(23.6) | 64(19.9) |
| Female | 134(83.7) | 123(76.4) | 257(80.1) |
| Education level of caregiver |  |  |  |
| High school or less | 114(71.3) | 117(72.7) | 231(71.0) |
| College or more | 46(28.7) | 44(27.3) | 90(28.0) |
| Marital status of caregiver |  |  |  |
| Unmarried | 14(8.8) | 23(14.3) | 37(11.5) |
| Married | 134(83.7) | 124(77.0) | 258(80.4) |
| Divorced or widowed | 12(7.5) | 14(8.7) | 26(8.1) |
| Household income levels† |  |  |  |
| ≤11,611 USD/year | 110(68.8) | 123(76.4) | 233(72.6) |
| > 11,611 USD/year | 50(31.2) | 38(23.6) | 88(27.4) |
| Awareness of HPV vaccines |  |  |  |
| Yes | 146(91.3) | 140(87.0) | 286(89.1) |
| No | 14(8.7) | 21(13.0) | 35(10.9) |
| Whether anyone in the same household has ever been diagnosed with HPV or cervical cancer |  |  |  |
| Have been diagnosed | 11(6.9) | 4(2.5) | 15(4.7) |
| Have not been diagnosed | 141(88.1) | 150(93.2) | 291(90.6) |
| Not clear | 8(5.0) | 7(4.3) | 15(4.7) |
| Study sites |  |  |  |
| Site A | 40(25.0) | 43(26.7) | 83(25.9) |
| Site B | 44(27.5) | 39(24.2) | 83(25.9) |
| Site C | 38(23.8) | 39(24.2) | 77(24.0) |
| Site D | 38(23.8) | 40(24.9) | 78(24.4) |

Data are mean (SD) or n (%). †The average household income in rural Sichuan Province was around RMB 80,000 in 2021 according to Sichuan Statistical Yearbook 2022.
1USD=6.89RMB. HPV = human papilloma virus.

A total of 32.1% (103/321) of participants scheduled appointments, with 42.2% (68/161) in the pay-it-forward arm, and 21.9% (35/160) in the standard-of-care arm (Fig. 1; appendix table 1, p 11). During the follow-up period, a total of 25.9% (83/321) received the vaccination, with 34.2% (55/161) in the pay-it-forward arm, and 17.5% (28/160) in the standard-of-care arm (appendix video, p2). Pay-it-forward participants were significantly more likely to receive a vaccination than those in the standard-of-care arm, with an adjusted proportional difference of 17.9% (95% CI: 8.7, 27.0%, p<0.001) (Fig 2). No severe adverse events within 24 hours following the vaccination were reported.

Pre-specified subgroup analyses (Fig. 2) showed that pay-it-forward was associated with a significant increase in vaccine uptake among girls 1) when female caregivers were present for the intervention (adjusted proportion difference= 20.2%, 9.7, 30.7%, p<0.001), 2) among caregivers aged 40 or above (adjusted proportional difference=18.4%, 7.6, 29.2%, p=0.001), 3) across education levels (high school or below: adjusted proportion difference=16.3%, 4.9, 27.7%, p=0.007; college or above: adjusted proportion difference=28.4%, 10.7, 46.2%, p=0.002), 4) among caregivers with lower household incomes (adjusted proportion difference=19.8%, 8.9-30.1%, p<0.001), 5) among caregivers who were aware of HPV vaccines before participating this study(adjusted proportion difference=20.2%, 10.9, 29.5%, p<0.001), 6) across two study sites (Site A: adjusted proportion difference=adjusted proportion difference=22.8%, 6.7-38.9%, p=0.006; Site C: adjusted proportion difference=25.4%, 4.3, 46.6%, p=0.029).

Among 83 girls who received the first dose vaccine, 69 of them received the second dose, with the second-dose vaccine uptake rates being 89.1% (49/55) and 71.4% (20/28) respectively in the pay-it-forward and standard-of-care arms (Fig. 3). Our analysis of vaccine confidence (Fig. 3) showed that pay-it-forward participants reported significantly higher levels of confidence in vaccine importance, safety, and effectiveness, for both nonavalent and bivalent HPV vaccines than those in the standard-of-care arm. A lower proportion of participants in the pay-it-forward arm showed vaccine delay compared to that in the standard-of-care arm (adjusted proportion difference = −18.4%, −27.6, −9.2%, p< 0.001; Fig. 3).

The total financial cost for the healthcare provider of implementing an HPV vaccination intervention for participants was $8,240 for the standard-of-care arm, and $12,676 for the pay-it-forward arm. Among participants in the pay-it-forward arm who received vaccination, 70.9% of participants (39/55) donated money, and the total donation amount was $332, with a median of $2.9 (IQR 1.6-9.6). The donation distribution by study site is shown in table 2 and appendix figure 5, p 9. The financial cost per person vaccinated was $294 in the standard-of-care arm and $230 in the pay-it-forward arm. The proportion of financial cost related to variable cost was 36.7% and 42.7% in the standard-of-care and pay-it-forward arm, respectively (appendix figure 6, p 9).

**Table 2.** Donations by study sites in the pay-it-forward arm (USD)

|  | Vaccinated | Donated | Total donation | Median donation | Minimum donation | Maximum donation | P value* |
| --- | --- | --- | --- | --- | --- | --- | --- |
| Site A (n=83) | 11 | 11(100.0) | 134.6 | 8.7 (5.1, 19.2) | 2.4 | 29.0. | 0.082 |
| Site B (n=83) | 18 | 16(88.9) | 160.6 | 5.1 (1.5, 14.9) | 0.1 | 47.9 | .. |
| Site C (n=77) | 17 | 8(47.1) | 13.5 | 2.3 (1.5, 3.3) | 1.5 | 7.3 | .. |
| Site D (n=78) | 9 | 4(44.4) | 23.5 | 2.9 (2.3, 4.0.) | 0.4 | 7.3 | .. |
| Total | 55 | 39(70.9) | 332.2 | 2.9 (1.6, 9.6) | 0.1 | 47.9 | .. |

Data are n, n(%) or median (IQR). \*Independent-Samples Kruskal-Wallis Test was used to compare differences of median donation amount between different sites. 1USD=6.89RMB.

Out of 55 girls vaccinated in the pay-it-forward arm, 37 (67.3%) girls wrote a postcard message to encourage others to vaccinate (appendix figure 7, pp 9-10). Out of the total of 37 messages, 18.9% (7/37) encouraged other girls to vaccinate, and 83.8% (31/37) expressed their wish for a world free of diseases and conveyed their blessings to other girls in the format of a text message or a poem.

## Discussion

HPV vaccination is the most effective way to prevent cervical cancer, but community-engaged strategies to promote uptake are limited. Our data demonstrated that the pay-it-forward strategy can increase HPV vaccine uptake compared to the standard-of-care. The strategy was also associated with greater vaccine confidence, reduced vaccine delay, generated community messages and engagement, and comparable costs. Our study extends the literature by focusing on an innovative incentive strategy for HPV vaccination, and leveraging prosocial tendencies to enhance public health. This is a rare example of a community-engaged intervention to reduce HPV vaccine delay and enhance vaccine uptake in a middle-income country context.

The pay-it-forward arm had a greater vaccine uptake rate than the standard-of-care. This is consistent with previous studies using pay-it-forward to improve influenza vaccination and other health services uptake.[18–20] The vaccination rate in the pay-it-forward arm was significantly higher than that among girls aged ≤18 years old in a 2021 national survey (12.4%).[8] Pay-it-forward also had a higher uptake compared to the impact of a 2015 UK-based financial incentive intervention study on HPV vaccine uptake (28.4%).[30] Although the UK-based study was old, China’s HPV vaccine services are about 10 years behind the UK, therefore these figures were presumably comparable. Pay-it-forward was also twice as effective among caregivers with lower household incomes compared to those with higher incomes, which might have implications for reducing HPV vaccination inequalities. The effect may be because of community-contributed financial support, locally relevant postcard messages, community engagement, or some combination.[31] Future research can be done to better understand how pay-it-forward motivated our participants to vaccinate so as to inform tailored strategies to address low vaccine uptake in similar contexts.

Pay-it-forward may be a novel incentive strategy and had lower costs per person vaccinated compared to the standard-of-care approach. Among those recruited in the pay-it-forward arm, most participants who received a vaccine voluntarily donated to support another girl. A systematic review suggested that conventional financial incentives (e.g., cash payments, vouchers) are largely effective in increasing vaccine uptake,[32] and our research adds to the literature by demonstrating the effectiveness of an innovative community-engaged incentive design. Despite small mean donations collected through the pay-it-forward strategy, we observed higher average donations in the two better-off regions than in the lower-income regions, suggesting the potential of creating a subsidization mechanism to support HPV vaccination in poorer regions. Of note, pay-it-forward will not replace the government role and should be understood as an incentive structure or complementary model to support future catch-up vaccination programs.

The pay-it-forward intervention generated additional community-created messages, mostly written by adolescent girls, to encourage future’s vaccination. Community engagement is essential to the success of health service programs. Community engagement focused on cultivating kindness may improve confidence, better acceptance of vaccine services, and our pay-it-forward participants showed significantly better vaccine confidence and less vaccine delay. Previous data showed that pay-it-forward was associated with increased community solidarity,[33] and vaccine confidence.[18] A systematic review demonstrated that pro-social interventions are associated with improved health outcomes for both givers and recipients.[31] Pay-it-forward may also provide a novel nudging approach for effective messaging and vaccine communication by fostering a culture of mutual support and trust.[31] This may be particularly relevant to regions where free or subsidized vaccine services are available, but have sub-optimal uptake rates due to vaccine hesitancy.

The study has some limitations. First, our study was implemented after the Chengdu municipal government rolled out the subsidized HPV vaccination program for 13-14-year-old girls, which might have improved public awareness, and created additional peripheral demand among other age groups for HPV vaccines. However, all participants were randomized, and we expect this change had a similar impact on our study outcomes between the two arms.

Second, our study had a clinic attendance rate of 54%. Our study was conducted during COVID-19 control measures, and there were COVID-19 cases and short-term (3-7 days) community-scale lockdowns to contain viral spread. This might have reduced participants’ likelihood to visit clinics for recruitment or not showing up for appointments, resulting in underestimated response and vaccination rates in the study. Finally, the study is conducted in a western province in China and the study sites were selected based on the availability of vaccines, organizational willingness, and capacity to collaborate. Selection bias caused by convenience sampling of study sites is possible, and generalizability to the country should be made cautiously.[34] But our study sites from different economic settings reflected common pathways for HPV vaccination in China.

Our study has important implications for practice, research and policy in China and other LMICs. First, this is a rare community-engaged intervention to increase HPV vaccine uptake among priority populations in a middle-income country. The research findings may have important implications for pro-social interventions to improve public trust and address vaccine delay. Second, the community engagement methods to generate contextually appropriate intervention materials and successful implementation of a novel multi-component strategy in clinical settings may support evidence-based practices and effective public messaging. Third, pay-it-forward is an innovative incentive design that leverages human kindness to expand public health services and address intention-to-behavior gap. The model is highly adaptable to local contexts where giving and reciprocity culture exist, and gift components can be contextualized.[31] Fourth, it can complement government-led HPV vaccination initiatives to broaden access for more individuals and might be a transition model to universal health coverage for catch-up HPV vaccination programs. Future research can explore potential mechanisms of how pay-it-forward works, the sustainability and scalability in similar contexts.

## Acknowledgments

The Bill & Melinda Gates Foundation (INV-034554, INV-003174), US NIH (NIAID R01AI158826), Nanjing Medical University Career Development Grant (NMUR20230008),, Jiangsu Provincial Professorship Career Development Grant (KY103R202309), and National Natural Science Foundation of China (82473742) funded this study. DH and WFG from Gates provided vaccine-related expertise and relevant contexts, but did not involve in the study design, implementation, data collection or data analysis. The authors are fully responsible for the content of this manuscript, and the views and opinions described in the publication reflect solely those of the authors. The authors are grateful to all participants. We would like to thank all collaborative health staff in Yulin Community Health Service Center, Longtan Community Health Service Center, Xinjin District Maternal and Child Healthcare Hospital, and Third People’s Hospital of Chengdu Eastern New Area for their contribution to the execution of this study. We thank all advisory group members who provided advice on contextual background.

## Contributors

DW and JL conceived the idea, designed the study and oversaw the project. SLT, JDT, WT and FY provided expert advice on the local contexts and study design throughout the project. HD and WFG provided vaccine-related expertise. YH coordinated local resources for fieldwork, and YFL, CYQ, YJL, YY implemented the study and collected data. YFL, CYQ and HDL cleaned the dataset, analyzed data and generated the figures and tables, and WT and FZ provided epidemiological and statistical expertise. YWX and JJO conducted economic analyses. JL, YFL, CYQ, YWX and DW wrote the draft. HJL, MJ, LL, JSS, GE, DX and all other co-authors provided constructive feedback on the manuscript. All authors reviewed and approved the final version of the manuscript.

## Data availability

Anonymous data with code book and data analysis code used in the main analysis are available in the Supporting Information files.

## Supporting information

➢ Video abstract
➢ Appendix Figures

Appendix Figure 1: Overview of pay-it-forward
Appendix Figure 2: Study site recruitment over time
Appendix Figure 3: Community engaged postcards
Appendix Figure 4: Educational pamphlet
Appendix Figure 5: Percentages of donations of vaccine costs by study sites
Appendix Figure 6. Breakdown of financial costs by category in proportions in the two arms
Appendix Figure 7: Translated example hand-written postcard messages from the participants
➢ Community engagement activities
➢ Telephone recruitment script
➢ Appendix tables

Appendix Table 1: Recruitment, appointment, and reasons for non-presence for vaccination
➢ Cost analysis section

Appendix Table 2. Cost calculations (in 2022 USD) for the study
Appendix Table 3. Outcome analyses for costs of each group
➢ Protocol

Original Trial Protocol (Version 1.0)
Protocol Version 3.0, 4 May, 2023
Amendments to the protocol
Protocol version history
➢ Data analysis code (R version 4.2.1)
➢ Questionnaire and codebook
➢ Consort checklist

